# Urinary iodine concentration after household use of iodine-fortified bouillon among women of reproductive age and preschool children in northern Ghana: A secondary analysis of the CoMIT trial

**DOI:** 10.64898/2026.07.07.26357449

**Authors:** Felix Kwaku Kyereh, Agartha N. Ohemeng, Reina Engle-Stone, K. Ryan Wessells, Sika M. Kumordzie, Charles D. Arnold, Jennie N. Davis, Emily R. Becher, Ahmed D. Fuseini, Kania W. Nyaaba, Xiuping Tan, Stephen A. Vosti, Seth Adu-Afarwuah

## Abstract

**Background:** Monitoring urinary iodine concentration (UIC) after sustained exposure informs iodine delivery strategies. We aimed to compare endline UIC among women of reproductive age (WRA) and preschool children (PSC) receiving multiple micronutrient-fortified (MMF) vs. iodine-only bouillon, and to describe population iodine status at baseline and endline.

**Methods:** This pre-specified secondary outcome analysis used Condiment Micronutrient Innovation Trial (CoMIT) data. Non-pregnant, non-lactating WRA (15–49 years) and PSC (2–5 years) were enrolled at the household level. Households received MMF or iodine-only bouillon for 9 months (38 weeks); both contained iodine (KIO[) at 30 µg/g. Baseline and endline UIC (µg/L) were measured by modified Sandell–Kolthoff reaction in spot urine samples. ANCOVA models, adjusted for baseline log-UIC and recruitment site, compared endline geometric mean UIC between trial arms. Median UIC at both timepoints was compared to WHO cut-offs (µg/L): <100 (insufficient), 100–199 (adequate), 200–299 (above requirements), and ≥300 (excessive); the prevalence of UIC <50 µg/L was calculated.

**Results:** Endline UIC data were available for 611 WRA and 630 PSC. Adjusted endline UIC did not differ by arm (geometric mean ratio [95% CI]: WRA 0.97 [0.86–1.10]; PSC 0.96 [0.84–1.10]). With arms combined, median UIC increased among WRA (100.5 to 124.6 µg/L) and PSC (109.6 to 136.9 µg/L), with fewer samples <50 µg/L at endline.

**Conclusions:** Adjusted endline UIC did not differ between trial arms. After 9 months’ use of iodine-fortified bouillon, median UIC remained adequate among WRA and PSC, with fewer samples <50 µg/L at endline.

**Trial registration:** ClinicalTrials.gov: NCT05178407; Pan-African Clinical Trial Registry: PACTR202206868437931.

## Introduction

Iodine is essential for thyroid hormone synthesis and for growth, neurodevelopment and metabolic homeostasis across the life course (Delić & Karanović Štambuk, 2026). Inadequate iodine intake during pregnancy and early childhood is associated with impaired cognitive development, suboptimal linear growth and adverse maternal health outcomes; therefore, women of reproductive age (WRA) and preschool children (PSC) are priority groups for iodine-status surveillance (Kampouri et al., 2023; Lin et al., 2025; Zimmermann, 2009).

Universal salt iodisation has reduced iodine deficiency disorders globally, yet iodine inadequacy persists in many low- and middle-income countries where the coverage, quality or use of iodised salt is inconsistent (Lin et al., 2025; Sáez-Ramírez et al., 2024). In sub-Saharan Africa, iodine status varies widely because of differences in food systems, household salt use and access to fortified foods (Lin et al., 2025; Zimmermann & Andersson, 2021). The World Health Organization (WHO) defines adequately iodised household salt as containing 15–40 ppm iodine and indicates that household access should preferably exceed 90% (World Health Organization, n.d.). Ghana has had salt iodisation policies for more than two decades, but the 2022 Ghana Demographic and Health Survey reported that 79.8% of households with tested salt had iodised salt, leaving approximately one-fifth without salt that tested positive for iodisation (Ghana Statistical Service [GSS] & ICF, 2024). However, iodised salt coverage does not necessarily ensure adequate iodine exposure because iodine concentrations may be insufficient, decline during storage or cooking, or be consumed in amounts below physiological requirements (Ghana Statistical Service [GSS] & ICF, 2024; United Nations Children’s Fund, 2023). Previous studies indicate that iodine exposure remains uneven in Ghana, with evidence of suboptimal iodine status among vulnerable women and young children in northern Ghana (Abu et al., 2019; Kubuga et al., 2019).

Bouillon cubes are widely used in household cooking across West Africa, including Ghana (Engle-Stone, Kumordzie, et al., 2024). Their widespread consumption among women and young children makes them a potential complementary vehicle for micronutrient fortification. The Condiment Micronutrient Innovation Trial (CoMIT) evaluated multiple micronutrient-fortified bouillon cubes containing vitamin A, vitamin B12, folate, iron, zinc, and iodine, compared with control bouillon cubes fortified with iodine only. Both formulations contained iodine as potassium iodate at the same concentration (30 µg/g bouillon) (Engle-Stone, Wessells, et al., 2024). The additional micronutrients included in the MMF bouillon formulation (i.e., iron, vitamin A and zinc) may be relevant to iodine physiology (Zimmermann & Andersson, 2021; Zimmermann & Köhrle, 2002). Iron is required for the haem-dependent enzyme thyroid peroxidase, and iron deficiency can impair thyroid hormone synthesis and blunt the response to iodine supplementation, whereas iron repletion may restore thyroidal responsiveness to iodine (Zimmermann & Köhrle, 2002). In populations with concurrent vitamin A and iodine deficiency, vitamin A deficiency may remove the suppressive effect of vitamin A on pituitary TSHβ expression, leading to elevated thyroid-stimulating hormone and increased thyroid size; vitamin A supplementation can reduce this thyroidal hyperstimulation (Zimmermann, 2007). Zinc also supports the deiodinase enzymes involved in the conversion of thyroxine to triiodothyronine (Severo et al., 2019).

Although iodine-containing co-fortified salt has been evaluated, it remains unknown whether co-fortifying bouillon with iodine plus iron, vitamin A and zinc alters iodine status compared with iodine fortification alone. It is also unclear whether replacing usual commercial bouillon with iodine-fortified study bouillon shifts population urinary iodine concentration (UIC) distributions where universal salt iodisation is established and commercial bouillon may already contribute to background iodine exposure. In this prespecified secondary outcome analysis of CoMIT, we compared endline UIC among WRA and PSC whose households received MMF or iodine-only bouillon and described population iodine status from baseline to endline using WHO cut-offs (WHO/UNICEF/ICCIDD, 2007).

## Key messages

- Among women of reproductive age and preschool children, endline UIC did not differ between the multiple micronutrient-fortified bouillon and iodine-only bouillon arms, indicating that added micronutrients did not measurably alter UIC when iodine content was the same.
- With trial arms combined, population median UIC was within the World Health Organization range for adequate iodine intake at baseline and endline, and the proportion of samples with UIC <50 µg/L was lower after 9 months of household use of iodine-fortified bouillon.
- Iodine-fortified bouillon may serve as an additional iodine delivery vehicle in settings where bouillon is widely consumed, but continued surveillance is needed to maintain adequacy and avoid excessive iodine exposure.

## Methods

### Study design and trial overview

The study design, setting, and procedures are described in detail elsewhere (Engle-Stone, Wessells, et al., 2024). Briefly, this was a community-based, randomised, controlled, double-masked trial conducted across 16 urban, peri-urban and rural sites in the Tolon and Kumbungu districts of the Northern Region of Ghana. The trial was registered on ClinicalTrials.gov (NCT05178407) and the Pan-African Clinical Trial Registry (PACTR202206868437931). The randomised comparison is reported in accordance with the CONSORT 2025 Statement and relevant CONSORT extensions, with the completed checklist provided as **Supplementary File 1**. No formal patient or public advisory group contributed to the trial design, analysis, or reporting; however, community entry and engagement were undertaken to support culturally appropriate recruitment, communication, and trial conduct.

No important changes to the trial methods, outcomes, or planned analyses were made after trial commencement. The present secondary analysis included 762 WRA and 662 PSC with at least one valid UIC measurement at baseline or endline. Before baseline assessment, all participants completed a three-week observational period. During this period, fieldworkers conducted up to three household visits to collect data on participant characteristics, dietary intake, morbidity, and any bouillon use.

Following completion of the observation period, eligible participants attended a mobile biospecimen collection site for the baseline visit where biological samples and other baseline measurements were collected and final eligibility for randomisation was confirmed. Households with participants who were found eligible were then randomly assigned to receive either MMF-fortified or iodine-only bouillon. Enrolled households received biweekly in-home visits, during which study-provided bouillon cubes were provided and data were collected on adherence and morbidity.

Two endline assessments in the parent trial were conducted after approximately nine months of exposure (35 and 38 weeks) for WRA and PSC.

### Participant eligibility, recruitment, and consent

Eligible participants were non-pregnant, non-lactating WRA (15–49 years) and preschool children aged 2–5 years residing in eligible households within the study districts. Each household contributed at most one participant per physiological group. Households were eligible if they were willing to use the study-provided bouillon cubes for the duration of the trial and had no household member enrolled in another study, requiring frequent blood transfusions for a chronic condition, or reporting an allergy to any ingredients contained in bouillon. Individuals were excluded if they had a severe acute or chronic illness, were unable or unwilling to provide informed consent, or did not plan to remain in the study area for the full duration of follow-up. Additional physiological-group-specific eligibility criteria, including biochemical, anthropometric, and morbidity thresholds, were applied as specified in the study protocol (Engle-Stone, Wessells, et al., 2024).

Recruitment began on January 15, 2023, and ended in May 2023; participant follow-up was completed on May 31, 2024. Recruitment was conducted through door-to-door household identification using a random-walk approach within predefined rural, peri-urban, and urban sampling areas (Flynn et al., 2013). Recruitment and eligibility screening were performed by trained enumerators fluent in English and Dagbani (the local dialect) using standardized questionnaires and procedures.

Written informed consent was obtained from participating WRA and from caregivers of participating preschool children, with consent documented by signature or thumbprint. Participants aged 15–17 years who had never married and were living with their parents provided assent, with caregiver consent; married participants aged 15–17 years provided their own informed consent. For non-literate participants, consent was obtained in the presence of an impartial witness who was not affiliated with the study. Ethical approval for the parent CoMIT trial was obtained from the Ghana Health Service Ethics Review Committee (GHS-ERC: 024/11/21) and the University of California, Davis Institutional Review Board (IRB ID: 1837253). Participants were considered enrolled only after completion of the baseline visit where confirmation of full eligibility and randomization to a study group occurred.

### Randomisation and blinding

Households, irrespective of the number of eligible physiological populations within their household, were the unit of randomisation. The trial statistician (C.D.A.) generated the allocation sequence using computerised block randomisation with variable block sizes of four or eight. Allocation was concealed using opaque envelopes prepared by study staff not involved in randomisation. After baseline procedures and confirmation of continued eligibility, designated trained study staff selected the four uppermost envelopes from the allocation stack of envelopes, and the participant selected one envelope to reveal their household study assignment. The assigned study code was then linked to the household and participant identifiers.

All bouillon cubes were identically packaged and labelled only with the study code and an associated group colour. Participants and field staff were aware of the study code but blinded to the micronutrient composition. Independent custodians at collaborating academic institutions maintained the code–formulation mapping. Randomisation codes were stored in sealed envelopes at secure sites, and pre-specified unblinding procedures were defined in the protocol (Engle-Stone, Wessells, et al., 2024).

### Intervention and adherence

Intervention households received bouillon cubes fortified with vitamin A, vitamin B12, folate, iron, zinc, and iodine, while iodine-only households received bouillon cubes fortified with iodine only. Both formulations contained iodine as potassium iodate (KIO[) at 30 μg/g bouillon. Assuming consumption of 2.0–2.5 g bouillon/day among WRA and ∼1 g/day among PSC, this provided approximately 60–75 μg/day of iodine to WRA and ∼30 μg/day to PSC. Thus, the two formulations were designed to provide equivalent iodine exposure from the study bouillon, with the MMF formulation differing from the iodine-only by the addition of vitamin A, folic acid, vitamin B12, iron, and zinc. The micronutrient composition of the study formulations and estimated daily doses are shown in **Supplementary Table S1**

Study bouillon cubes were delivered to households every two weeks by trained enumerators, with household rations based on household size and estimated bouillon consumption of 2.5 g/capita/day. Households were instructed to prepare meals as usual, substituting study-provided bouillon cubes for commercial products. Adherence was monitored biweekly using inventories of remaining study and commercial bouillon cubes, self-reported use, and structured questionnaires on purchases, usage patterns, and sharing. Households with persistently low adherence to study-bouillon use were identified but were not restricted from using commercial bouillon. Potential contamination was minimised by household-level randomisation and provision of a single bouillon formulation per household. Adherence was monitored for trial implementation purposes but was not used to define the analytical population or included as a covariate in the present intention-to-treat analyses.

### Data collection and outcome measurement

Following recruitment and preliminary eligibility screening, participant- and household-level characteristics were collected at household visits during the approximately 3-week observation period before the baseline visit. Structured questionnaires were administered by trained enumerators to collect participant- and household-level characteristics, including household head sex, age, education, and occupation. Socio-economic status was assessed using a household asset index (Filmer & Pritchett, 2001), and household food security was measured using the Household Food Insecurity Access Scale (HFIAS) according to FANTA-3 guidelines (Coates et al., 2007). Household water and sanitation were assessed using standardized questionnaires and classified as improved or unimproved according to World Health Organization/UNICEF Joint Monitoring Programme definitions (World Health Organization, 2021). These pre-randomisation variables were considered as potential covariates in the analysis.

At the end of the observation period, eligible participants were invited to a mobile biospecimen collection site for the baseline visit, where spot midstream urine samples were collected from WRA and PSC before randomisation. The outcome of interest in this secondary analysis was urinary iodine concentration (UIC, µg/L), a prespecified secondary outcome of the parent CoMIT trial. Two endline assessments were conducted 35 and 38 weeks following the baseline visit, after approximately nine months of exposure to the study bouillon. The present analysis used UIC from the 38-week endline assessment as the endline outcome.

Following collection, urine samples were aliquoted into cryovials, stored in a battery-operated cold box maintained at 4–8°C, and transported daily to the District Hospital at Tolon, where they were stored at −86°C with regular temperature monitoring. Samples were subsequently transported to the University of Ghana for longer-term frozen storage before analysis. UIC was determined at the Iodine Deficiency Disorders Laboratory, University of Ghana, Accra, using a modified Sandell–Kolthoff reaction (Global Alliance for Improved Nutrition (GAIN), 2015). Baseline and endline samples from the same participant were analysed on the same day. Samples were analysed in duplicate, and the mean value was used in the analysis. Each analytical batch included working iodine standards, duplicate standards, and internal urine controls. Results were accepted only when internal quality-control values were within the laboratory’s predefined acceptable range. In brief, samples were digested with ammonium persulfate at 95°C for 60 minutes, absorbance was read at 405 nm, and iodine concentrations were determined from a standard curve. Laboratory analysts were blinded to treatment allocation. Potential intervention-related harms were assessed systematically during the biweekly household visits for bouillon delivery. At each visit, field staff asked households whether any household member had experienced any complaint perceived to be related to the study bouillon; reported concerns were recorded and reviewed by the study team.

### Sample size and statistical analysis

The parent trial was powered to detect a standardised mean difference of 0.26 with 80% power at a two-sided α of 0.05, requiring 234 participants per trial arm in each physiological population, or 468 WRA and 468 PSC across the two trial arms. Although the calculation was based on a two-sample comparison, the prespecified ANCOVA models adjusted for baseline outcome values, which was expected to improve precision. In the present analysis, valid endline UIC data were available for 611 WRA and 630 PSC. Cross-sectional summaries used all valid UIC values at each time point. Baseline–endline descriptive transition analyses and ANCOVA treatment-effect models were restricted to participants with valid UIC at both time points; because model covariates were complete, these analyses included 595 WRA and 610 PSC.

Analyses were conducted separately for WRA and PSC using R version 4.6.0. Participant and household characteristics were summarised descriptively for each population group. Analyses followed the published trial protocol, which specified adjustment for recruitment site as a fixed-effect covariate. Because WRA and PSC were analysed in separate population-specific models, and each household contributed at most one participant to each model, a household-level random effect was not fitted. Recruitment site was retained in all ANCOVA models to account for the protocol-specified site structure.

UIC was right-skewed; therefore, baseline and endline UIC values were natural log-transformed before regression analysis. Model-based results are presented as geometric means, geometric mean ratios and 95% confidence intervals, obtained by back-transforming estimates from the log scale. UIC values outside the laboratory-specified analytical plausibility range (<5 or >2000 µg/L) were coded as missing: 13 among WRA (9 baseline, 4 endline) and 11 among PSC (7 baseline, 4 endline), all below 5 µg/L.

The primary trial-arm comparison analysed participants according to randomised assignment among those with valid baseline and endline UIC data. In the minimally adjusted ANCOVA model, log-transformed endline UIC was modelled with randomised trial arm, baseline log-UIC and recruitment site included as fixed effects. Adjusted geometric mean endline UIC values and geometric mean ratios comparing the MMF bouillon arm with the iodine-only bouillon arm were estimated from marginal means on the log scale and back-transformed.

A fully adjusted ANCOVA model was fitted as a secondary analysis to assess robustness and improve precision; the minimally adjusted ANCOVA model remained primary for inference. Prespecified candidate covariates were WRA age and educational level, household head education, household size, HFIAS, household water insecurity, drinking water source and sanitation facility for WRA; and household head education, household size, household water insecurity, drinking water source, sanitation facility, child age and child sex for PSC. Candidate covariates were retained using partial F tests at P < 0.20. The final fully adjusted models retained WRA educational level, household sanitation facility and household water insecurity for WRA, and child age and sex for PSC.

Effect modification by baseline UIC was assessed by adding a randomised trial arm × baseline log-UIC interaction term to the fully adjusted ANCOVA model. Interaction tests were evaluated at P < 0.10. For descriptive presentation, stratified estimates were also calculated by baseline UIC category, defined as below versus at or above the within-population median baseline UIC.

Because both study bouillon formulations contained iodine at 30 µg/g, baseline–endline changes in iodine status were summarised descriptively with trial arms pooled and were not interpreted as treatment effects. Cross-sectional summaries included median UIC and WHO population-level UIC categories: <100 µg/L (insufficient), 100–199 µg/L (adequate), 200–299 µg/L (above requirements) and ≥300 µg/L (excessive). Distributional proportions of spot urine samples with UIC <50 µg/L, <100 µg/L and ≥300 µg/L were calculated with Wilson 95% confidence intervals. Selected UIC percentiles were summarised at baseline and endline by population cohort, with 95% confidence intervals estimated using non-parametric bootstrap resampling with 10,000 resamples within each population-by-time point stratum. For both WRA and PSC, these categories, proportions and percentiles were interpreted as population-level programmatic summaries, not as diagnostic thresholds or prevalence estimates of individual iodine deficiency or excess. This distinction was particularly relevant for PSC because the WHO UIC categories are less directly age-specific for this population (WHO/UNICEF/ICCIDD, 2007; World Health Organization, 2013).

Because endline UIC data were missing for 151 of 762 WRA (19.8%) and 32 of 662 PSC (4.8%), a multiple-imputation sensitivity analysis was conducted among WRA to assess the robustness of the primary complete-case treatment-effect estimate. Twenty imputed datasets were generated by chained equations under a missing-at-random assumption. The imputation model included all variables in the minimally adjusted model, together with the prespecified WRA covariates. Estimates were pooled using Rubin’s rules. Multiple imputation was not performed for PSC because endline UIC missingness was low.

All statistical tests were two-sided. The primary treatment effect was evaluated at α = 0.05. Interaction tests were evaluated at α = 0.10, and covariate screening used P < 0.20. Model assumptions were assessed using residual-versus-fitted plots, normal Q–Q plots of standardised residuals and Cook’s distance.

## Results

### Participant flow and analysis population

Participant flow is shown in **Figure 1**. Of 769 WRA and 670 PSC randomised in the parent CoMIT trial, 762 WRA and 662 PSC had at least one valid UIC measurement and were eligible for this secondary analysis; 7 WRA and 8 PSC had no valid UIC measurement at either visit. Baseline and endline UIC were available for 746 and 611 WRA, respectively, and 642 and 630 PSC, respectively. Endline UIC was unavailable for 151 WRA and 32 PSC, with reasons detailed in Figure 1. The ANCOVA sample, requiring valid baseline and endline UIC plus complete covariate data, included 595 WRA and 610 PSC; fully adjusted models used the same denominators.

**Figure 1.**
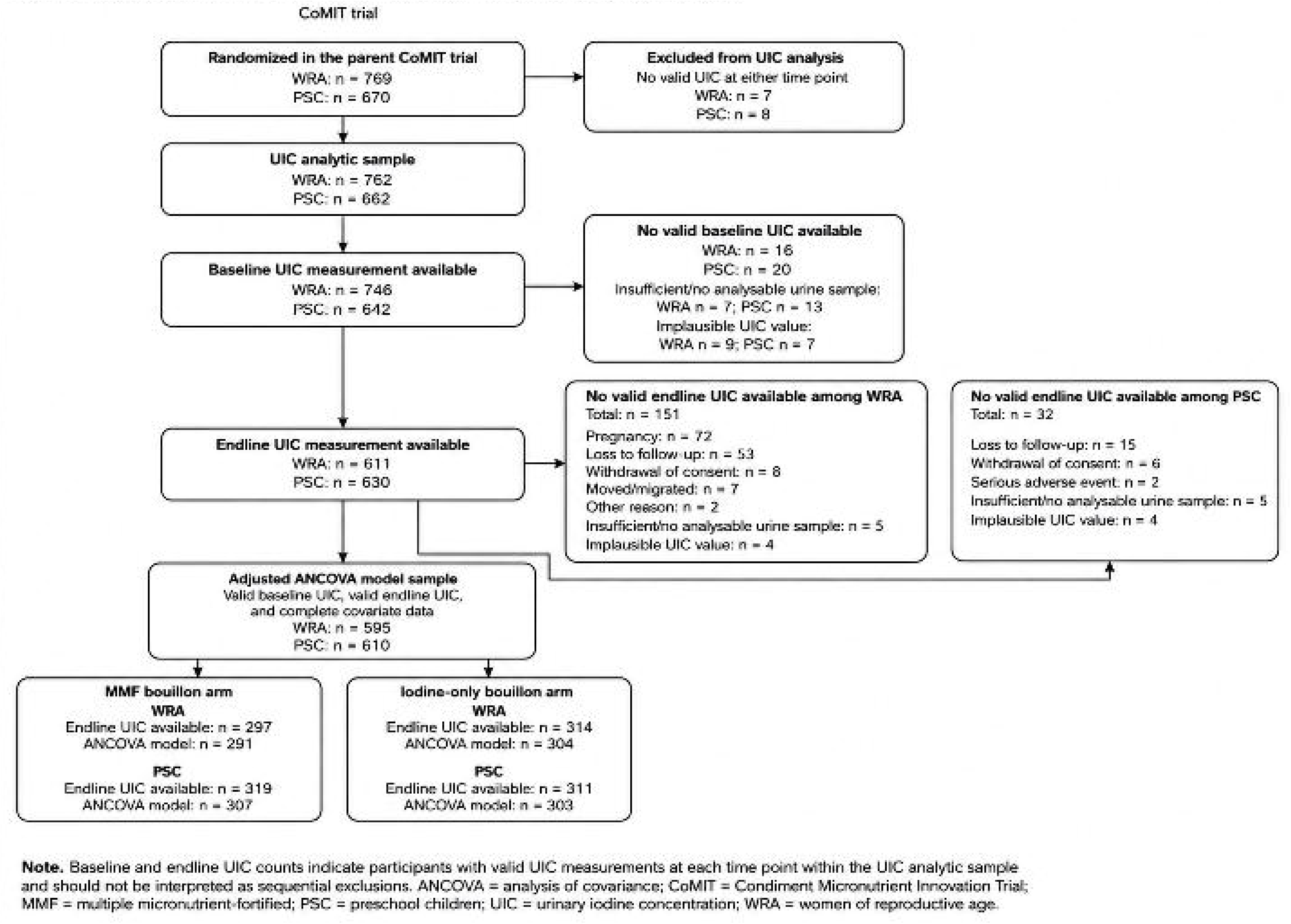
Participant flow for the urinary iodine concentration analysis among women of reproductive age and preschool children in the CoMIT trial. Baseline and endline UIC counts indicate participants with valid UIC measurements at each time point within the UIC analytic sample and should not be interpreted as sequential exclusions. The adjusted ANCOVA sample included participants with valid baseline UIC, valid endline UIC and complete covariate data.

### Baseline characteristics

Baseline household and individual characteristics of the analytic sample are shown in **Table 1**. Among WRA, the mean age was 33.9 years, and less than one-fifth had attained at least secondary education. Among PSC, the mean age was 41.2 months, and approximately one-half were female.

**Table 1.** Baseline characteristics of women of reproductive age and preschool children participating in the Condiment Micronutrient Innovation Trial in northern Ghana.

| Variable | Women of reproductive age (WRA)<br>(n = 762) | Preschool children (PSC)<br>(n = 662) |
| --- | --- | --- |
| **Household level** |  |  |
| District (Tolon) | 382/762 (50.1%) | 322/662 (48.6%) |
| Residence (rural) | 376/762 (49.3%) | 335/662 (50.6%) |
| Highest formal educational level of household head ≥ Secondary (yes/no) | 96/755 (12.7%) | 84/657 (12.8%) |
| Household Food Insecurity Access Scale (HFIAS) score | 3.2 ± 3.4 (762) | 3.0 ± 3.3 (662) |
| HFIAS categorised |  |  |
| Food secure | 310/762 (40.7%) | 276/662 (41.7%) |
| Mild food insecure | 133/762 (17.5%) | 104/662 (15.7%) |
| Moderate food insecure | 260/762 (34.1%) | 217/662 (32.8%) |
| Severe food insecure | 59/762 (7.7%) | 65/662 (9.8%) |
| Household size | 11.8 ± 6.1 (762) | 11.6 ± 5.9 (662) |
| Drinking water source (improved) | 404/762 (53.0%) | 356/662 (53.8%) |
| Sanitation (improved) | 97/762 (12.7%) | 79/662 (11.9%) |
| **Individual level** |  |  |
| Age | 33.9 ± 9.6 years | 41.2 ± 10.1 months |
| Educational level ≥ Secondary (yes/no) – women only | 139/749 (18.6%) | - |
| Sex (female) |  | 332/662 (50.2%) |

**Notes:**
Values are presented as mean ± standard deviation (n) for continuous variables and n/N (%) for categorical variables. Denominators vary across variables due to missing data. Age is presented in years for women of reproductive age and in months for preschool children. Educational level ≥ secondary refers to women only. Improved drinking water source and improved sanitation were defined according to WHO/UNICEF Joint Monitoring Programme classifications. HFIAS, Household Food Insecurity Access Scale. For binary variables, values reflect the proportion in the indicated category.

Food insecurity was common in both analytic populations, affecting 59.3% of women’s households and 58.3% of children’s households. Moderate or severe food insecurity was reported in 41.8% and 42.6% for WRA and PSC households, respectively. Baseline characteristics for each analysis population are shown in Supplementary Tables S2a and S2b. No intervention-related adverse effects were reported.

### Urinary iodine concentration by trial arm

Urinary iodine concentration by trial arm is presented in **Table 2**. Among WRA, median baseline UIC was 101.8 µg/L (IQR 57.4–168.3) in the MMF bouillon group and 99.6 µg/L (IQR 57.0–179.2) in the iodine-only bouillon arm. At endline, median UIC was 125.0 µg/L (IQR 68.8–193.8) in the MMF bouillon arm and 123.9 µg/L (IQR 73.7–191.7) in the iodine-only bouillon arm. In the minimally adjusted model adjusting for baseline log-UIC and recruitment site, there was no evidence of a difference between trial arms (geometric mean ratio [GMR] 0.97; 95% CI 0.86–1.10; P = 0.67). Results were similar in the secondary analysis adjusting additionally for WRA educational level, household sanitation, and household water insecurity (GMR 0.99; 95% CI 0.88–1.12; P = 0.85).

**Table 2.** Urinary iodine concentration of women of reproductive age and preschool children at baseline and endline, by trial arm^1^.

| Urinary iodine concentration,<br>µg/L | Women of reproductive age |  |  | Preschool children |  |  |
| --- | --- | --- | --- | --- | --- | --- |
|  | MMF bouillon | Iodine-only | *p*-value | MMF bouillon | Iodine-only | *p*-value |
| Baseline, n | 377 | 369 |  | 321 | 321 |  |
| Baseline, median (Q1, Q3)<sup>2</sup> | 101.8 (57.4, 168.3) | 99.6 (57.0, 179.2) |  | 108.8 (58.3, 191.1) | 110.8 (69.4, 196.8) |  |
| Endline, n | 297 | 314 |  | 319 | 311 |  |
| Endline, median (Q1, Q3)<sup>2</sup> | 125.0 (68.8, 193.8) | 123.9 (73.7, 191.7) |  | 134.0 (80.8, 234.0) | 140.1 (84.9, 253.8) |  |
| Adjusted model sample, n<sup>3</sup> | 291 | 304 |  | 307 | 303 |  |
| Baseline- and site-adjusted geometric mean UIC at endline (95% CI), primary model<sup>4</sup> | 110 (99, 122) | 113 (103, 125) | 0.67 | 131 (118, 146) | 137 (123, 152) | 0.57 |
| Geometric mean ratio (95% CI), MMF vs Iodine-only<sup>5</sup> | 0.97 (0.86, 1.10) | Reference | 0.67 | 0.96 (0.84, 1.10) | Reference | 0.57 |

<sup>1</sup>MMF bouillon = multiple micronutrient-fortified bouillon cube containing vitamin A, folic acid, vitamin B12, iron, zinc, and iodine; Iodine-only = bouillon cube fortified with iodine only. WRA = women of reproductive age; PSC = preschool children; UIC = urinary iodine concentration.
<sup>2</sup>Values are median (Q1, Q3), where Q1 and Q3 denote the first and third quartiles, respectively. n denotes the number of participants with valid UIC data at the corresponding time point. Baseline and endline descriptive summaries are based on participants with valid UIC data at the corresponding time point.
<sup>3</sup>Adjusted model sample includes participants with valid baseline UIC, valid endline UIC, and complete data for the covariates included in the primary adjusted model. Sample sizes may differ across baseline descriptive summaries, endline descriptive summaries, and adjusted model estimates because UIC availability and covariate completeness varied by time point and analysis.
<sup>4</sup>Values are geometric means (95% CI) obtained by back-transforming model-estimated mean log-UIC from an ANCOVA model adjusted for baseline log-UIC and recruitment site (primary model).
<sup>5</sup>Geometric mean ratio (95% CI) for MMF bouillon versus iodine-only, derived from the same ANCOVA model. A GMR of 1.00 indicates no difference between trial arms; values below 1.00 indicate lower endline UIC in the MMF bouillon arm relative to the iodine-only bouillon arm. P-values are from the test of the treatment effect in the adjusted model.

Among PSC, median baseline UIC was 108.8 µg/L (IQR 58.3–191.1) in the MMF bouillon arm and 110.8 µg/L (IQR 69.4–196.8) in the iodine-only bouillon arm. At endline, median UIC was 134.0 µg/L (IQR 80.8–234.0) in the MMF bouillon arm and 140.1 µg/L (IQR 84.9–253.8) in the iodine-only bouillon arm. In the minimally analysis adjusting for baseline log-UIC and recruitment site, there was no evidence of a difference between trial arms (GMR 0.96; 95% CI 0.84–1.10; P = 0.57). Results were similar in the secondary analysis adjusting additionally for child age and sex (GMR 0.98; 95% CI 0.85–1.12; P = 0.72).

Baseline iodine status did not modify the association between trial arm and endline UIC among WRA (p-interaction = 0.75) or PSC (p-interaction = 0.96) (**Supplementary Table S3**).

### Population iodine status over time

Population iodine status over time is shown in **Table 3**. Analyses were combined across trial arms because both study formulations contained iodine at the same concentration. Among WRA, median UIC increased from 100.5 µg/L at baseline to 124.6 µg/L at endline, with both values within the WHO range for adequate population iodine status. The proportion of WRA with UIC <50 µg/L decreased from 21.8% (95% CI 19.0–25.0) at baseline to 13.9% (95% CI: 11.4–16.9) at endline. The proportion with UIC <100 µg/L decreased from 49.7% (95% CI: 46.2–53.3) to 39.9% (95% CI: 36.1–43.9), whereas the proportion with UIC ≥300 µg/L increased from 4.8% (95% CI: 3.5–6.6) to 9.0% (95% CI: 7.0–11.5).

**Table 3.**
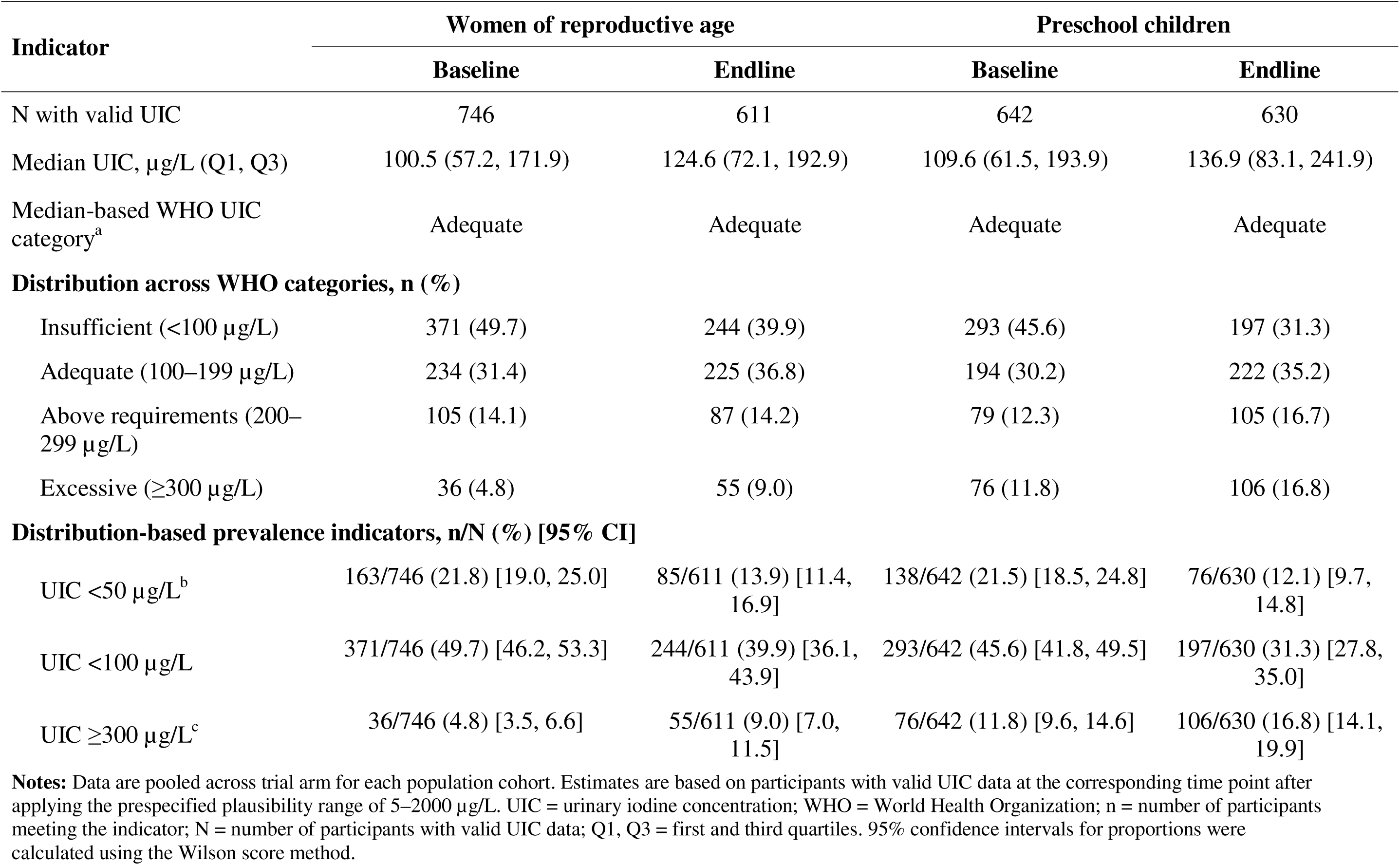
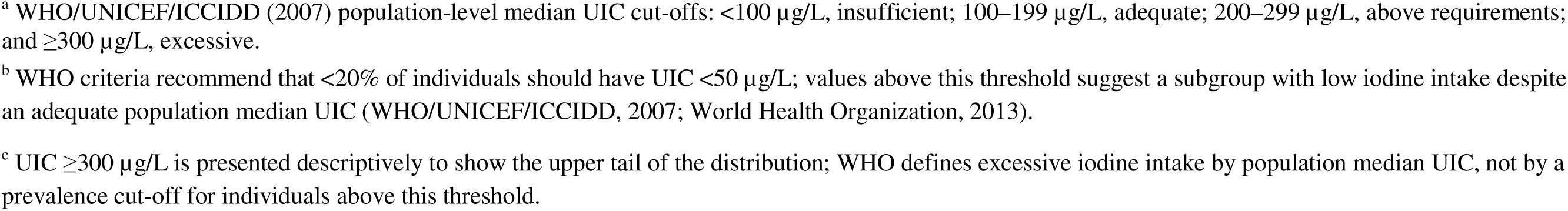
Population-level urinary iodine status indicators among women of reproductive age and preschool children at baseline and endline.

| Indicator | Women of reproductive age |  | Preschool children |  |
| --- | --- | --- | --- | --- |
|  | Baseline | Endline | Baseline | Endline |
| N with valid UIC | 746 | 611 | 642 | 630 |
| Median UIC, µg/L (Q1, Q3) | 100.5 (57.2, 171.9) | 124.6 (72.1, 192.9) | 109.6 (61.5, 193.9) | 136.9 (83.1, 241.9) |
| Median-based WHO UIC category<sup>a</sup> | Adequate | Adequate | Adequate | Adequate |
| **Distribution across WHO categories, n (%)** |  |  |  |  |
| Insufficient (<100 µg/L) | 371 (49.7) | 244 (39.9) | 293 (45.6) | 197 (31.3) |
| Adequate (100–199 µg/L) | 234 (31.4) | 225 (36.8) | 194 (30.2) | 222 (35.2) |
| Above requirements (200–299 µg/L) | 105 (14.1) | 87 (14.2) | 79 (12.3) | 105 (16.7) |
| Excessive (≥300 µg/L) | 36 (4.8) | 55 (9.0) | 76 (11.8) | 106 (16.8) |
| **Distribution-based prevalence indicators, n/N (%) [95% CI]** |  |  |  |  |
| UIC <50 µg/L<sup>b</sup> | 163/746 (21.8) [19.0, 25.0] | 85/611 (13.9) [11.4, 16.9] | 138/642 (21.5) [18.5, 24.8] | 76/630 (12.1) [9.7, 14.8] |
| UIC <100 µg/L | 371/746 (49.7) [46.2, 53.3] | 244/611 (39.9) [36.1, 43.9] | 293/642 (45.6) [41.8, 49.5] | 197/630 (31.3) [27.8, 35.0] |
| UIC ≥300 µg/L<sup>c</sup> | 36/746 (4.8) [3.5, 6.6] | 55/611 (9.0) [7.0, 11.5] | 76/642 (11.8) [9.6, 14.6] | 106/630 (16.8) [14.1, 19.9] |

**Notes:** Data are pooled across trial arm for each population cohort. Estimates are based on participants with valid UIC data at the corresponding time point after applying the prespecified plausibility range of 5–2000 µg/L. UIC = urinary iodine concentration; WHO = World Health Organization; n = number of participants meeting the indicator; N = number of participants with valid UIC data; Q1, Q3 = first and third quartiles. 95% confidence intervals for proportions were calculated using the Wilson score method.
<sup>a</sup> WHO/UNICEF/ICCIDD (2007) population-level median UIC cut-offs: <100 µg/L, insufficient; 100–199 µg/L, adequate; 200–299 µg/L, above requirements; and ≥300 µg/L, excessive.
<sup>b</sup> WHO criteria recommend that <20% of individuals should have UIC <50 µg/L; values above this threshold suggest a subgroup with low iodine intake despite an adequate population median UIC (WHO/UNICEF/ICCIDD, 2007; World Health Organization, 2013).
<sup>c</sup> UIC ≥300 µg/L is presented descriptively to show the upper tail of the distribution; WHO defines excessive iodine intake by population median UIC, not by a prevalence cut-off for individuals above this threshold.

Among PSC, median UIC increased from 109.6 µg/L at baseline to 136.9 µg/L at endline, remaining within the WHO adequate range at both time points. The proportion of PSC with UIC <50 µg/L decreased from 21.5% (95% CI: 18.5–24.8) at baseline to 12.1% (95% CI: 9.7–14.8) at endline. The proportion with UIC <100 µg/L decreased from 45.6% (95% CI: 41.8–49.5) to 31.3% (95% CI: 27.8–35.0), whereas the proportion with UIC ≥300 µg/L increased from 11.8% (95% CI: 9.6–14.6) to 16.8% (95% CI: 14.1–19.9). Selected percentiles of the UIC distribution at each time point, with bootstrap 95% confidence intervals, are provided in **Supplementary Table S4**.

### Distributional changes in paired analyses

Kernel density plots showed a rightward shift in the UIC distribution from baseline to endline in both populations, consistent with the observed reduction in the proportion with UIC <50 µg/L and increase in the proportion with UIC ≥300 µg/L (**Figure 2**). In paired analyses, among participants with baseline UIC <50 µg/L, 79.5% of WRA (95% CI: 71.9–85.5) and 83.6% of PSC (95% CI: 76.2–89.0) had UIC ≥50 µg/L at endline. The full pattern of within-participant movement across the four WHO UIC categories and across the <50 and ≥300 µg/L thresholds is shown in **Supplementary Tables S5 and S6.**

**Figure 2.**
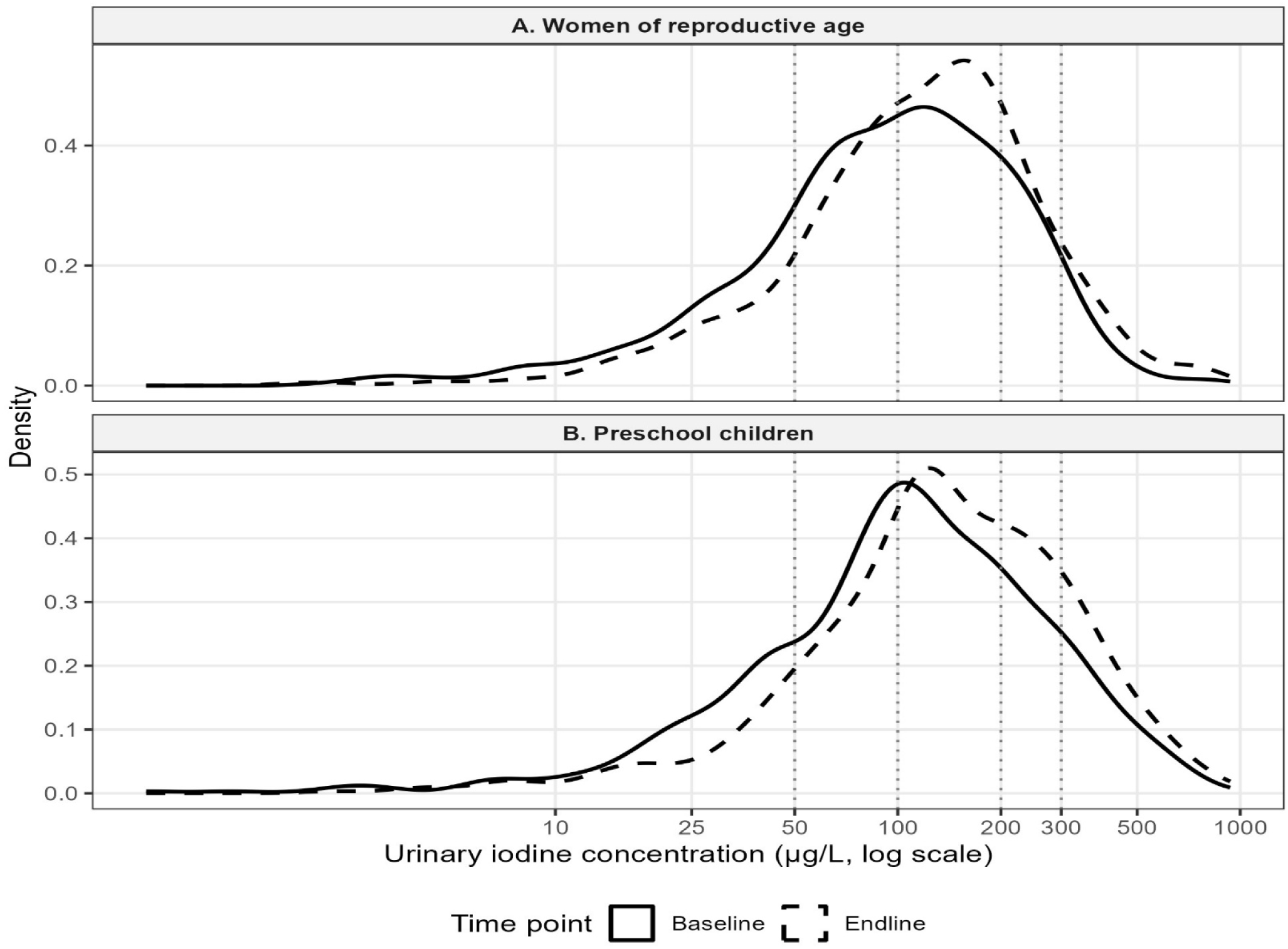
Distribution of urinary iodine concentration at baseline and endline among women of reproductive age and preschool children. Kernel density plots show log-transformed urinary iodine concentration, with trial arms combined, for women of reproductive age (A) and preschool children (B). Solid lines indicate baseline distributions and dashed lines indicate endline distributions. Vertical dotted lines mark 50, 100, 200, and 300 µg/L. In both populations, the distribution shifted to the right from baseline to endline, consistent with fewer observations in the very low UIC range and a small increase in the upper tail beyond 300 µg/L.

### Sensitivity analyses

A multiple imputation sensitivity analysis was conducted among WRA to assess the potential influence of missing endline UIC data for the treatment effect model. In the full WRA cohort used for the imputation model, endline UIC data were missing for 19.8% of WRA. After imputation using chained equations with 20 imputed datasets, the pooled GMR for endline UIC comparing the MMF bouillon arm with the iodine-only bouillon arm was 0.98 (95% CI: 0.87–1.09), consistent with the complete-case primary analysis.

## Discussion

In this community-based, randomised, controlled, double-masked trial in northern Ghana, adjusted endline UIC did not differ between the MMF-fortified and iodine-only bouillon arms among WRA or PSC. Median UIC remained within the WHO population-level adequate range in both populations at baseline and endline and increased descriptively over the trial period in both populations. These findings indicate that co-fortification with additional micronutrients did not produce a measurable difference in UIC beyond iodine-only fortification under the conditions of this trial.

The absence of a between-arm difference is consistent with the intervention contrast. Both bouillon formulations contained iodine at the same concentration and were delivered under similar household conditions; therefore, iodine exposure was not expected to differ substantially by treatment assignment (Engle-Stone, Wessells, et al., 2024). The trial therefore tested whether adding other micronutrients to iodine-fortified bouillon altered UIC, rather than whether iodine-fortified bouillon improved iodine status relative to non-iodine-fortified bouillon. Although iron, vitamin A and zinc are biologically relevant to thyroid hormone metabolism, their inclusion in the multiple-micronutrient formulation did not result in a detectable additional effect on UIC.

The pooled baseline-to-endline changes provide useful context for interpreting population iodine exposure during the trial. At baseline, median UIC was adequate in both WRA and PSC. However, more than one-fifth of spot samples in each population had UIC <50 µg/L, indicating that a substantial proportion of samples reflected very low recent iodine exposure. After approximately nine months of household exposure to iodine-fortified study bouillon, the proportion of samples with UIC <50 µg/L was below 20% in both populations. These descriptive changes are consistent with higher iodine exposure during the trial period, but they cannot be attributed solely to the study bouillon because both trial arms received iodine-containing bouillon and there was no non-iodine control group. Seasonal variation in diet or hydration, household iodised-salt use, commercial bouillon use, and broader changes in the iodine food supply may also have contributed.

These findings are consistent with evidence from iodised salt and fortified-condiment programmes in low- and middle-income countries, where iodine interventions can improve population iodine status but may not eliminate heterogeneity in intake (Knowles et al., 2023; Sáez-Ramírez et al., 2024; Zimmermann & Andersson, 2021). In Ghana, previous studies have documented persistent suboptimal iodine intake despite long-standing salt iodisation policies, particularly in rural and food-insecure settings (Coomson & Aryeetey, 2022; Simpong et al., 2016).

Although baseline median UIC was adequate in the present trial, the distribution of UIC suggested that some population subgroups remained vulnerable to low iodine exposure. This supports the rationale for considering centrally produced, widely consumed condiments such as bouillon cubes as complementary vehicles for iodine delivery, particularly where household salt iodisation coverage or quality is variable (Eilander et al., 2023; Garcia-Casal et al., 2016).

Several contextual factors may explain the observed shifts in the UIC distribution. Households were advised to replace commercial bouillon with the study-provided bouillon, except when study rations were exhausted. Because the iodine content of commercial bouillon was not measured in this trial, baseline-to-endline changes may reflect differences between study and commercial products, changes in total bouillon consumption, intermittent use of commercial bouillon, or iodine intake from other dietary sources. Previous work in northern Ghana found substantial but brand-variable iodine concentrations in commercial bouillon cubes, supporting their plausibility as an important background iodine source (Abizari et al., 2017). The regular household distribution of study bouillon may also have supported sustained exposure over the intervention period, reflecting habitual use of bouillon in household meals rather than a short-term supplementation model.

This study has several strengths. The double-masked randomised design reduced selection and performance bias, and the inclusion of both WRA and PSC strengthens the relevance of the findings for maternal and child nutrition. Frequent household follow-up and monitored use of the study-provided bouillon support internal validity. Iodine status was assessed using UIC, the recommended biomarker for population-level iodine assessment, and the analyses accounted for baseline UIC, recruitment site and the skewed distribution of UIC values (WHO/UNICEF/ICCIDD, 2007).

The findings should also be interpreted in light of several limitations. Spot UIC reflects recent iodine intake and is subject to substantial within-person variability; therefore, the proportions of samples with UIC <50 or ≥300 µg/L should be interpreted as descriptors of the population UIC distribution rather than precise estimates of individual iodine deficiency or excess. By design, both trial arms received iodine-fortified bouillon, so the trial could not estimate the absolute effect of iodine-fortified bouillon compared with non-iodine-fortified bouillon. Missing endline UIC data, particularly among women, may have reduced precision, although multiple-imputation analyses were consistent with complete-case findings. The trial did not assess thyroid function or longer-term functional outcomes, and iodine intake from non-bouillon sources, especially household iodised salt, was not separately quantified. Generalisability may therefore be greatest for settings with similar bouillon consumption patterns and comparable iodine-fortification contexts.

From a programme perspective, the findings add to evidence that iodine-fortified bouillon may complement existing iodine interventions in settings where iodine intake remains heterogeneous despite salt iodisation policies. However, fortified bouillon should not be viewed as a replacement for universal salt iodisation, salt-quality monitoring or salt-reduction strategies. This is particularly important in settings where discretionary salt intake is high (Davis et al., 2024). Although median UIC remained within the adequate range, the proportion of spot samples with UIC ≥300 µg/L increased from baseline to endline in WRA and PSC, from 4.8% to 9.0% and from 11.8% to 16.8%, respectively. These estimates may overstate habitual high iodine intake because of day-to-day variation in spot UIC, but they reinforce the need to monitor the full UIC distribution, including the upper tail, as fortified bouillon or other iodine vehicles are scaled up.

In conclusion, adjusted endline UIC did not differ between the MMF-fortified and iodine-only bouillon arms, suggesting no measurable additional effect of co-fortification on UIC under equivalent iodine provision. In pooled descriptive analyses, median UIC remained within the WHO adequate range and increased from baseline to endline among both WRA and PSC. Because both trail arms received iodine-fortified bouillon, the study cannot determine the effect of iodine-fortified bouillon relative to non-iodine-fortified bouillon. Continued population-level monitoring is needed to maintain iodine adequacy while avoiding excessive iodine exposure as fortified-condiment strategies evolve.

## Declarations

## Ethics approval and consent

Ethical approval for the parent CoMIT trial was obtained from the Ghana Health Service Ethics Review Committee (GHS-ERC: 024/11/21) and the University of California, Davis Institutional Review Board (IRB ID: 1837253). Written informed consent was obtained from adult participants and from parents or legal guardians of minors, with assent obtained from minors according to the approved study procedures. Married adolescents were consented according to the procedures approved by the ethics committees.

## Use of artificial intelligence

The authors used a generative artificial intelligence tool to support language editing and clarity of expression. The authors reviewed, revised and verified all AI-assisted text and take full responsibility for the accuracy, integrity and originality of the final manuscript.

## Data availability statement

The data that support the findings of this study are openly available in the Open Science Framework repository for the Condiment Micronutrient Innovation Trial (CoMIT) Project at https://doi.org/10.17605/OSF.IO/T3ZRN, reference number T3ZRN. The statistical analysis plan for the present analysis is available at https://osf.io/t3zrn/files/ktm46, and the data files supporting the analyses are available at https://osf.io/t3zrn/files/jhmra and https://osf.io/t3zrn/files/g5fq2.

## Author contributions

Felix Kwaku Kyereh: Formal analysis, methodology, writing—original draft, writing—review and editing.

Agartha N. Ohemeng: Supervision, writing—review and editing.

Reina Engle-Stone: Conceptualisation, funding acquisition, methodology, supervision, writing—review and editing.

K. Ryan Wessells: Methodology, writing—review and editing.

Sika M. Kumordzie: Investigation, methodology, project administration, writing—review and editing. Charles D. Arnold: Methodology, formal analysis, writing—review and editing.

Jennie N. Davis: Investigation, methodology, writing—review and editing. Emily R. Becher: Investigation, methodology, writing—review and editing.

Ahmed D. Fuseini: Investigation, project administration, writing—review and editing. Kania W. Nyaaba: Investigation, project administration, writing—review and editing. Xiuping Tan: Methodology, writing—review and editing.

Stephen A. Vosti: Conceptualisation, funding acquisition, writing—review and editing.

Seth Adu-Afarwuah: Conceptualisation, methodology, funding acquisition, supervision, writing—review and editing.

## Acknowledgements

The authors thank the study participants, field staff, laboratory personnel, community leaders, and district health authorities in Tolon and Kumbungu for their contributions to the CoMIT trial.

## References

Abizari, A. R., Dold, S., Kupka, R., & Zimmermann, M. B. (2017). More than two-thirds of dietary iodine in children in northern Ghana is obtained from bouillon cubes containing iodized salt. Public Health Nutrition, 20(6), 1107–1113. 10.1017/S1368980016003098

Abu, B. A. Z., Oldewage-Theron, W., & Aryeetey, R. N. O. (2019). Risks of excess iodine intake in Ghana: Current situation, challenges, and lessons for the future. Annals of the New York Academy of Sciences, 1446(1), 117–138. 10.1111/nyas.13988

Coates, J., Swindale, A., & Bilinsky, P. (2007). Household Food Insecurity Access Scale (HFIAS) for measurement of household food access: Indicator guide (v. 3). Food and Nutrition Technical Assistance Project, Academy for Educational Development. https://www.fao.org/fileadmin/user_upload/eufao-fsi4dm/doc-training/hfias.pdf

Coomson, J. B., & Aryeetey, R. N. O. (2022). Scoping review of diet-related health outcomes and associated risk factors in Ghana. African Journal of Food, Agriculture, Nutrition and Development, 22(2), 19496–19524. 10.18697/ajfand.107.21795

Davis, J. N., Kumordzie, S. M., Arnold, C. D., Wessells, K. R., Nyaaba, K. W., Adams, K. P., Tan, X., Becher, E. R., Vosti, S. A., Adu-Afarwuah, S., & Engle-Stone, R. (2024). Consumption of discretionary salt and salt from bouillon among households, women, and young children in Northern Region, Ghana: A mixed-methods study with the Condiment Micronutrient Innovation Trial (CoMIT) Project. Current Developments in Nutrition, 8(3), Article 102088. 10.1016/j.cdnut.2024.102088

Delić, T., & Karanović Štambuk, S. (2026). Iodine in health and disease: A comprehensive review. Nutrients, 18(8), Article 1262. 10.3390/nu18081262

Eilander, A., Verbakel, M. R., & Dötsch-Klerk, M. (2023). The potential of condiments, seasonings, and bouillon cubes to deliver essential micronutrients in Asia: Scenario analyses of iodine and iron fortification. Nutrients, 15(3), Article 616. 10.3390/nu15030616

Engle-Stone, R., Kumordzie, S. M., Luo, H., Wessells, K. R., Adu-Afarwuah, S., Njebayi, A., Teta, I., Régis, Y.-L., Gyimah, E., Vosti, S. A., & Adams, K. P. (2024). The potential for bouillon fortification to reduce dietary micronutrient inadequacy: Modeling analyses using national survey data from Cameroon, Ghana, and Haiti. Current Developments in Nutrition, 8(11), Article 104485. 10.1016/j.cdnut.2024.104485

Engle-Stone, R., Wessells, K. R., Haskell, M. J., Kumordzie, S. M., Arnold, C. D., Davis, J. N., Becher, E. R., Fuseini, A. D., Nyaaba, K. W., Tan, X., Adams, K. P., Lietz, G., Vosti, S. A., & Adu-Afarwuah, S. (2024). Effect of multiple micronutrient-fortified bouillon on micronutrient status among women and children in the Northern Region of Ghana: Protocol for the Condiment Micronutrient Innovation Trial (CoMIT), a community-based randomized controlled trial. PLOS ONE, 19(5), Article e0302968. 10.1371/journal.pone.0302968

Filmer, D., & Pritchett, L. H. (2001). Estimating wealth effects without expenditure data—or tears: An application to educational enrollments in states of India. Demography, 38(1), 115–132. 10.1353/dem.2001.0003

Flynn, A., Tremblay, P. F., Rehm, J., & Wells, S. (2013). A modified random walk door-to-door recruitment strategy for collecting social and biological data relating to mental health, substance use, addiction, and violence problems in a Canadian community. International Journal of Alcohol and Drug Research, 2(2), 7–16. 10.7895/ijadr.v2i2.143

Garcia-Casal, M. N., Peña-Rosas, J. P., Mclean, M., De-Regil, L. M., Zamora, G., & Consultation Working Groups. (2016). Fortification of condiments with micronutrients in public health: From proof of concept to scaling up. Annals of the New York Academy of Sciences, 1379(1), 38–47. 10.1111/nyas.13185

Ghana Statistical Service, & ICF. (2024). Ghana Demographic and Health Survey 2022. https://dhsprogram.com/pubs/pdf/FR387/FR387.pdf

Global Alliance for Improved Nutrition, Ghana Health Service, & United Nations Children’s Fund. (2017). National iodine survey report: Ghana 2015. https://www.unicef.org/ghana/media/1296/file/UN735926.pdf

Kampouri, M., Tofail, F., Rahman, S. M., Gustin, K., Vahter, M., & Kippler, M. (2023). Gestational and childhood urinary iodine concentrations and children’s cognitive function in a longitudinal mother-child cohort in rural Bangladesh. International Journal of Epidemiology, 52(1), 144–155. 10.1093/ije/dyac110

Knowles, J., Codling, K., Houston, R., & Gorstein, J. (2023). Introduction to the programme guidance for the use of iodised salt in processed foods and its pilot implementation, strengthening strategies to improve iodine status. PLOS ONE, 18(10), Article e0274301. 10.1371/journal.pone.0274301

Kubuga, C. K., Abizari, A. R., & Song, W. O. (2019). Iodine status of reproductive age women and their toddlers in northern Ghana improved through household supply of iodized salt and weekly indigenous meal consumption. PLOS ONE, 14(5), Article e0216931. 10.1371/journal.pone.0216931

Lin, J., Tan, H. L., & Ge, H. (2025). Global, regional, and national burden of iodine deficiency in reproductive women from 1990 to 2019, and projections to 2035: A systematic analysis for the Global Burden of Disease Study in 2019. International Journal of Women’s Health, 17, 1863–1875. 10.2147/IJWH.S513856

Sáez-Ramírez, D. M., Chacon-Torrico, H., & Hernández-Vásquez, A. (2024). Household consumption of adequately iodized salt: A multi-country analysis of socioeconomic disparities. Nutrients, 16(21), Article 3787. 10.3390/nu16213787

Severo, J. S., Morais, J. B. S., de Freitas, T. E. C., Andrade, A. L. P., Feitosa, M. M., Fontenelle, L. C., de Oliveira, A. R. S., Cruz, K. J. C., & do Nascimento Marreiro, D. (2019). The role of zinc in thyroid hormones metabolism. International Journal for Vitamin and Nutrition Research, 89(1–2), 80–88. 10.1024/0300-9831/a000262

Simpong, D. L., Adu, P., Bashiru, R., Morna, M. T., Yeboah, F. A., Akakpo, K., & Ephraim, R. K. (2016). Assessment of iodine status among pregnant women in a rural community in Ghana: A cross-sectional study. Archives of Public Health, 74, Article 8. 10.1186/s13690-016-0119-y

United Nations Children’s Fund. (2023). Iodine. Retrieved June 19, 2026, from https://data.unicef.org/topic/nutrition/iodine/

World Health Organization. (n.d.). Households consuming adequately iodized salt (≥15 parts per million). Retrieved May 31, 2026, from https://www.who.int/data/nutrition/nlis/info/households-consuming-adequately-iodized-salt-%28-15-parts-per-million%29

World Health Organization. (2013). Urinary iodine concentrations for determining iodine status in populations (WHO/NMH/NHD/EPG/13.1). https://www.who.int/publications/i/item/WHO-NMH-NHD-EPG-13.1

World Health Organization, & United Nations Children’s Fund. (2021). Progress on household drinking water, sanitation and hygiene 2000–2020: Five years into the SDGs. World Health Organization. https://www.who.int/publications/i/item/9789240030848

World Health Organization, United Nations Children’s Fund, & International Council for the Control of Iodine Deficiency Disorders. (2007). Assessment of iodine deficiency disorders and monitoring their elimination: A guide for programme managers (3rd ed.). World Health Organization. https://www.who.int/publications/i/item/9789241595827

Zimmermann, M. B. (2007). Interactions of vitamin A and iodine deficiencies: Effects on the pituitary-thyroid axis. International Journal for Vitamin and Nutrition Research, 77(3), 236–240. 10.1024/0300-9831.77.3.236

Zimmermann, M. B. (2009). Iodine deficiency. Endocrine Reviews, 30(4), 376–408. 10.1210/er.2009-0011

Zimmermann, M. B., & Andersson, M. (2021). Global endocrinology: Global perspectives in endocrinology: Coverage of iodized salt programs and iodine status in 2020. European Journal of Endocrinology, 185(1), R13–R21. 10.1530/EJE-21-0171

Zimmermann, M. B., & Köhrle, J. (2002). The impact of iron and selenium deficiencies on iodine and thyroid metabolism: Biochemistry and relevance to public health. Thyroid, 12(10), 867–878. 10.1089/105072502761016494

