## Supplementary material for "Urinary iodine concentration after household use of iodine-fortified bouillon among women of reproductive age and preschool children in northern Ghana: A secondary analysis of the CoMIT trial": Supplementary_Materials_iodine_manuscript.docx

| **Supplementary Table S1. Micronutrient composition of study bouillon formulations and estimated daily micronutrient doses** | | | | |
| --- | --- | --- | --- | --- |
| **Micronutrient (fortificant)** | **MMF bouillon concentration** | **Iodine-only concentration** | **Estimated daily dose for WRA** | **Estimated daily dose for PSC** |
| Vitamin A (retinyl palmitate) | 200 μg RE/g | Not added | 400–500 μg RE/day | 200 μg RE/day |
| Folic acid | 80 μg/g | Not added | 160–200 μg/day | 80 μg/day |
| Vitamin B12 | 1.2 μg/g | Not added | 2.4–3.0 μg/day | 1.2 μg/day |
| Iron (FePP/CA/TSC) | 4 mg/g | Not added | 8–10 mg/day | 4 mg/day |
| Zinc (ZnO) | 3 mg/g | Not added | 6–7.5 mg/day | 3 mg/day |
| Iodine (KIO₃) | 30 μg/g | 30 μg/g | 60–75 μg/day | 30 μg/day |
| **Note:** Values represent the active form of each nutrient and do not include overage. Estimated daily doses assume bouillon consumption of 2.0–2.5 g/day among WRA and 1 g/day among PSC, as specified in the parent trial protocol. WRA, women of reproductive age; PSC, preschool children; MMF, multiple micronutrient-fortified; RE, retinol equivalents; FePP/CA/TSC, ferric pyrophosphate/citric acid/trisodium citrate; ZnO, zinc oxide; KIO₃, potassium iodate. | | | | |

**Supplementary Table S2. Background characteristics of the study participants**

**Supplementary Table S2a. Baseline Characteristics of Non-pregnant, Non-lactating Women of Reproductive Age by Study Arm**

|  | **Characteristic** | | **Overall (N=762)** | **MMF (n=383)** | **Iodine only (n=379)** |
| --- | --- | --- | --- | --- | --- |
| **Age, years**, mean (SD) | | | 33.9 (9.6) | 34.2 (9.3) | 33.5 (9.9) |
| **District, n (%)** | | |  |  |  |
| Kumbungu | | | 380 (49.9) | 192 (50.1) | 188 (49.6) |
| Tolon | | | 382 (50.1) | 191 (49.9) | 191 (50.4) |
| **Residence/stratum, n (%)** | | |  |  |  |
| Rural | | | 376 (49.3) | 184 (48.0) | 192 (50.7) |
| Urban | | | 386 (50.7) | 199 (52.0) | 187 (49.3) |
| Household size, median (IQR) | | | 10.0 (7.2–14.0) | 10.0 (7.0–14.0) | 11.0 (8.0–15.0) |
| **Household water source, n (%)** | | |  |  |  |
| Improved | | | 404 (53.0) | 204 (53.3) | 200 (52.8) |
| Unimproved | | | 358 (47.0) | 179 (46.7) | 179 (47.2) |
| **Household sanitation facility, n (%)** | | |  |  |  |
| Improved | | | 97 (12.7) | 54 (14.1) | 43 (11.3) |
| Unimproved | | | 665 (87.3) | 329 (85.9) | 336 (88.7) |
| **Household head education, n (%)** | | |  |  |  |
| Below secondary | | | 659 (87.3) | 332 (87.4) | 327 (87.2) |
| Secondary or more | | | 96 (12.7) | 48 (12.6) | 48 (12.8) |
| **Household food insecurity category, n (%)** | | |  |  |  |
| Food secure | | | 310 (40.7) | 155 (40.5) | 155 (40.9) |
|  | | Mildly food insecure | 133 (17.5) | 71 (18.5) | 62 (16.4) |
| Moderately food insecure | | | 260 (34.1) | 125 (32.6) | 135 (35.6) |
| Severely food insecure | | | 59 (7.7) | 32 (8.4) | 27 (7.1) |
| HFIAS score, median (IQR) | | | 2.0 (0.0–6.0) | 2.0 (0.0–5.0) | 3.0 (0.0–6.0) |

Note. Values are n (%) unless otherwise indicated. UIC = urinary iodine concentration; HFIAS = Household Food Insecurity Access Scale; IQR = interquartile range; SD = standard deviation. Percentages are based on non-missing denominators. Missing values: baseline UIC n=7 (MMF=3, iodine only=4); household head education n=7 (MMF=3, iodine only=4).

**Table 2b. Baseline characteristics of preschool children by study arm**

| **Characteristic** | **Overall (N=662)** | **MMF (n=333)** | **Iodine only (n=329)** |
| --- | --- | --- | --- |
| Child age, months, mean (SD) | 41.2 (10.1) | 40.1 (10.0) | 42.4 (10.1) |
| **Sex, n (%)** |  |  |  |
| Female | 332 (50.2) | 171 (51.4) | 161 (48.9) |
| Male | 330 (49.8) | 162 (48.6) | 168 (51.1) |
| **District, n (%)** |  |  |  |
| Kumbungu | 340 (51.4) | 168 (50.5) | 172 (52.3) |
| Tolon | 322 (48.6) | 165 (49.5) | 157 (47.7) |
| **Residence/stratum, n (%)** |  |  |  |
| Rural | 335 (50.6) | 175 (52.6) | 160 (48.6) |
| Urban | 327 (49.4) | 158 (47.4) | 169 (51.4) |
| Household size, median (IQR) | 10.0 (7.0–14.0) | 10.0 (7.0–14.0) | 10.0 (8.0–15.0) |
| **Household water source, n (%)** |  |  |  |
| Improved | 356 (53.8) | 174 (52.3) | 182 (55.3) |
| Unimproved | 306 (46.2) | 159 (47.7) | 147 (44.7) |
| **Household sanitation facility, n (%)** |  |  |  |
| Improved | 79 (11.9) | 47 (14.1) | 32 (9.7) |
| Unimproved | 583 (88.1) | 286 (85.9) | 297 (90.3) |
| **Household head education, n (%)** |  |  |  |
| None or preschool | 513 (78.1) | 266 (80.4) | 247 (75.8) |
| Primary | 60 (9.1) | 30 (9.1) | 30 (9.2) |
| Secondary or more | 84 (12.8) | 35 (10.6) | 49 (15.0) |
| **Household food insecurity category, n (%)** |  |  |  |
| Food secure | 276 (41.7) | 148 (44.4) | 128 (38.9) |
| Mildly food insecure | 104 (15.7) | 52 (15.6) | 52 (15.8) |
| Moderately food insecure | 217 (32.8) | 102 (30.6) | 115 (35.0) |
| Severely food insecure | 65 (9.8) | 31 (9.3) | 34 (10.3) |
| HFIAS score, median (IQR) | 2.0 (0.0–5.0) | 2.0 (0.0–5.0) | 3.0 (0.0–6.0) |

Notes: Values are n (%) unless otherwise indicated. UIC = urinary iodine concentration; HFIAS = Household Food Insecurity Access Scale; IQR = interquartile range; SD = standard deviation. Percentages are based on non-missing denominators. Missing values: household head education n=5 (MMF=2, iodine only=3).

**Supplementary Table S3. Stratum-specific geometric mean ratios for endline urinary iodine concentration comparing the MMF bouillon group with the iodine-only bouillon arm, by baseline iodine status, among women of reproductive age and preschool children.**

| **Baseline UIC stratum** | **Women of reproductive age** | | **Preschool children** | |
| --- | --- | --- | --- | --- |
|  | **GMR (95% CI)ᵃ** | **Pᵇ** | **GMR (95% CI)ᵃ** | **Pᵇ** |
| ≥ median baseline UIC | 0.98 (0.82, 1.17) | 0.82 | 0.92 (0.76, 1.11) | 0.36 |
| < median baseline UIC | 1.00 (0.84, 1.18) | 0.98 | 1.03 (0.85, 1.24) | 0.79 |
| P-interaction^c^ | 0.75 | | 0.96 | |

**Notes:** Baseline UIC strata are defined by the within-population median baseline UIC. MMF bouillon = multiple micronutrient-fortified bouillon cube containing vitamin A, folic acid, vitamin B12, iron, zinc, and iodine; iodine-only = bouillon fortified with iodine only. UIC = urinary iodine concentration; GMR = geometric mean ratio (MMF bouillon vs. iodine-only).

^a^ Stratum-specific GMRs are descriptive. They were estimated from a covariate-adjusted ANCOVA model on natural-log-transformed endline UIC with a treatment × baseline-UIC-stratum interaction, adjusted for baseline log-UIC, recruitment site, and pre-specified covariates retained at P < 0.20 in screening (women of reproductive age: maternal education, household sanitation, household water insecurity; preschool children: child age, child sex).

^b^ Within-stratum P-values for the contrast of MMF bouillon vs. iodine-only.

^c^ P-interaction is the P-value for the treatment × continuous baseline log-UIC interaction term in the covariate-adjusted ANCOVA model and constitutes the formal test of effect modification by baseline iodine status.

| **Supplementary Table S4. Selected percentiles of UIC at baseline and endline, with bootstrap 95% confidence intervals** | | | | | | | | |
| --- | --- | --- | --- | --- | --- | --- | --- | --- |
| **Population** | **Time point** | **n** | **10th** | **25th** | **50th** | **75th** | **90th** | **95th** |
| WRA | Baseline | 746 | 28.5 (25.5, 33.2) | 57.2 (50.3, 62.2) | 100.5 (93.2, 109.1) | 171.9 (158.2, 183.4) | 253.4 (238.6, 269.1) | 295.7 (284.9, 315.7) |
| WRA | Endline | 611 | 38.4 (30.8, 46.7) | 72.1 (64.5, 79.1) | 124.6 (110.9, 133.6) | 192.9 (180.8, 204.3) | 292.6 (256.1, 315.1 | 368.0 (327.6, 422.9) |
| PSC | Baseline | 642 | 30.6 (25.4, 35.7) | 61.5 (52.5, 70.3) | 109.6 (101.0, 116.6) | 193.9 (183.5, 207.9) | 317.5 (294.2, 352.5) | 418.4 (373.5, 478.7) |
| PSC | Endline | 630 | 45.7 (39.0, 51.0) | 83.1 (75.0, 90.9) | 136.9 (126.7, 147.3) | 241.9 (224.6, 264.4) | 365.9 (338.5, 407.1) | 475.4 (422.2, 525.1) |
| **Notes:** Values are selected percentiles of urinary iodine concentration in µg/L, with bootstrap 95% confidence intervals shown in parentheses. Confidence intervals were estimated using non-parametric bootstrap resampling with 10,000 resamples within each population and time point. The n values represent participants with valid urinary iodine concentration measurements at each time point, after excluding missing values and values outside the prespecified plausible range (<5 or >2000 µg/L). WRA = women of reproductive age; PSC = preschool children; UIC = urinary iodine concentration. | | | | | | | | |

| **Supplementary Table S5. Paired transition matrix across WHO UIC categories among participants with valid baseline and endline UIC** | | | | |
| --- | --- | --- | --- | --- |
| **Women of reproductive age, n = 595** | | | | |
| **Baseline UIC category** | **Endline <100** | **Endline 100–199** | **Endline 200–299** | **Endline ≥300** |
| <100 | 150 (49.3%) | 105 (34.5%) | 30 (9.9%) | 19 (6.2%) |
| 100–199 | 61 (34.7%) | 69 (39.2%) | 28 (15.9%) | 18 (10.2%) |
| 200–299 | 21 (23.9%) | 34 (38.6%) | 20 (22.7%) | 13 (14.8%) |
| ≥300 | 4 (14.8%) | 11 (40.7%) | 8 (29.6%) | 4 (14.8%) |
| **Preschool children, n = 610** | | | | |
| **Baseline UIC category** | **Endline <100** | **Endline 100–199** | **Endline 200–299** | **Endline ≥300** |
| <100 | 115 (41.2%) | 95 (34.1%) | 32 (11.5%) | 37 (13.3%) |
| 100–199 | 50 (26.6%) | 70 (37.2%) | 36 (19.1%) | 32 (17.0%) |
| 200–299 | 16 (21.1%) | 30 (39.5%) | 16 (21.1%) | 14 (18.4%) |
| ≥300 | 9 (13.4%) | 23 (34.3%) | 16 (23.9%) | 19 (28.4%) |
| **Notes**: Data are pooled across trial arm for each population cohort and restricted to participants with valid UIC measurements at both baseline and endline. Row percentages are shown; therefore, each row sums to 100%, allowing interpretation of movement from each baseline UIC category to endline categories. UIC categories are based on concentration bands: <100, 100–199, 200–299, and ≥300 µg/L. | | | | |

| **Supplementary Table S6. Paired changes in lower-tail and upper-tail urinary iodine concentration among participants with valid baseline and endline measurements** | | |
| --- | --- | --- |
| **Indicator** | **Women of reproductive age, n = 595** | **Preschool children, n = 610** |
| Baseline UIC <50 µg/L | 132/595 (22.2%) [19.0, 25.7] | 128/610 (21.0%) [17.9, 24.4] |
| Endline UIC <50 µg/L | 82/595 (13.8%) [11.2, 16.8] | 73/610 (12.0%) [9.6, 14.8] |
| Baseline <50 to endline ≥50 µg/L | 105/132 (79.5%) [71.9, 85.5] | 107/128 (83.6%) [76.2, 89.0] |
| Baseline ≥50 to endline <50 µg/L | 55/463 (11.9%) [9.2, 15.1] | 52/482 (10.8%) [8.3, 13.9] |
| UIC <50 µg/L at both time points | 27/595 (4.5%) [3.1, 6.5] | 21/610 (3.4%) [2.3, 5.2] |
| Baseline UIC ≥300 µg/L | 27/595 (4.5%) [3.1, 6.5] | 67/610 (11.0%) [8.7, 13.7] |
| Endline UIC ≥300 µg/L | 54/595 (9.1%) [7.0, 11.7] | 102/610 (16.7%) [14.0, 19.9] |
| Baseline <300 to endline ≥300 µg/L | 50/568 (8.8%) [6.7, 11.4] | 83/543 (15.3%) [12.5, 18.6] |
| Baseline ≥300 to endline <300 µg/L | 23/27 (85.2%) [67.5, 94.1] | 48/67 (71.6%) [59.9, 81.0] |
| UIC ≥300 µg/L at both time points | 4/595 (0.7%) [0.3, 1.7] | 19/610 (3.1%) [2.0, 4.8] |
| **Notes**: Data are pooled across trial arm for each population cohort and restricted to participants with valid urinary iodine concentration measurements at both baseline and endline. Values are n/N (%) [95% CI]. Confidence intervals are Wilson score 95% CIs. For baseline and endline prevalence rows, the denominator is the total paired sample. For transition rows, the denominator is the number of participants in the relevant baseline category. Persistent low or high urinary iodine concentration rows are presented as proportions of the total paired sample. | | |
